# We have not registered any cholera outbreak following the vaccination in 2018 in the Albertine region. A resilience study of the Region’s journey towards the elimination of cholera outbreaks

**DOI:** 10.1101/2025.09.23.25336511

**Authors:** Nathan Tumuhamye, Lynn M. Atuyambe, Godfrey Bwire, Freddie Ssengooba, Simon Kasasa, Roy William Mayega

## Abstract

**Introduction:** World Health Assembly 2011 adopted a resolution on ‘strengthening national health emergency and resilience of health systems and following this resolution, an operational framework for climate resilient Health systems was formulated to guide health systems resilience analysis. Cholera has been endemic in Uganda for over five decades, with the Albertine region among the most affected. The highest burden was recorded before 2000, but cases have since declined after Uganda introduced integrated prevention measures like Oral Cholera Vaccination (OVC).

**Methods:** A qualitative study was conducted in Hoima, Kasese, and Kikuube districts. Data were collected through 13 key informant interviews with health officials and partners, 10 focus group discussions with community members, and 9 in-depth interviews with cholera survivors and caregivers, involving 87 participants in total. Participants were purposively selected based on their roles in cholera prevention and response. Analysis was done in two stages, first, the manifest content analysis followed by latent content analysis. Transcripts generated from the transcripts were read and re-read by three 3 selected research assistants, who then assigned codes (Open coding).

**Results:** Seven themes emerged as central to resilience to cholera shocks. OCV campaigns were seen as transformative, with districts reporting no outbreaks since. Epidemic preparedness and response planning enabled rapid mobilization, while health education improved community hygiene practices. Water and sanitation improvements, including protected sources and latrine coverage, reduced transmission. Rapid response systems and treatment centers minimized fatalities. Community mobilization through Village Health Teams enhanced surveillance and health education, while cross-border surveillance, though constrained, improved case detection.

**Conclusion:** Uganda’s declining cholera trend reflects synergies between vaccination, WASH, preparedness, and community engagement. Sustaining resilience requires periodic OCV booster campaigns, reinforcement of hygiene practices, improved cross-border coordination with neighbouring countries, permanent isolation facilities, and expanded safe water and sanitation access to consolidate progress toward cholera elimination.

## Background and introduction

Cholera outbreaks globally have been significantly linked to high rainfall, as the increased water flow can exacerbate the spread of the Vibrio cholerae bacteria (1–5). Heavy rainfall often leads to flooding, which can contaminate drinking water sources with sewage and other pollutants, creating ideal conditions for cholera transmission (6, 7). According to the World Health Organization (WHO), areas with poor sanitation infrastructure are particularly vulnerable during rainy seasons as the waterborne pathogen easily spreads through the faecal-oral route when water sources become compromised (8). This relationship between rainfall and cholera outbreaks has been observed in numerous regions, including parts of Sub-Saharan Africa and South Asia, where seasonal rains regularly trigger spikes in cholera cases (9). WHO has set a 2030 target that aims to reduce cholera deaths by 90% and eliminate transmission in at least 20 countries by 2030, underscoring the global commitment to addressing this threat (10).

Uganda has been hit by cholera for the last fifty-four years, with the first cases reported in 1971 by the World Health Organisation (11). The burden of cholera was quite high until 2000 when cases started decreasing (11–13) In the period between 2011 and 2016, Cholera accounted for 1,850 cases and 45 deaths, with Nebbi, Hoima, Buliisa, and Mbale contributing 60% of all reported cholera cases (14). In the same period, seventeen districts were responsible for 90% of all reported cholera cases in the country with greater Hoima, Kasese and Nebbi contributing 37% indicating that these districts are the most affected in Uganda. The Albertine region lies along the Lake Albert and River Nile in the mid-western end of the country, and the region comprises of the following districts: Nebbi and Zombo in the north, the greater Hoima and Kibale district in the west and Kasese district in the southwest. The region suffers recurrent floods and landslides due to heavy rains. These have far-reaching impacts on the affected areas escalating recurrent epidemics of cholera, malaria, and viral hemorrhagic fevers.

Uganda has implemented an integrated approach towards prevention and control of cholera with a focus on Water, sanitation and Hygiene (WASH), the implementation of Oral Cholera vaccination (OVC) in the most affected districts (15) and a robust surveillance, cases management and risk communication interventions (16). Since the introduction of OVC, districts that implemented this program have registered a significant decrease in cholera cases with some registering no case following the vaccination. While OVC is believed to have been the critical point in the elimination of cholera, it is important to note that over the years when Uganda was hit by cholera endemic, the ministry of health has implemented several initiatives geared towards the elimination of cholera and building the country’s resilience to disease outbreak. In this paper, we provide evidence of what communities affected by cholera as well as the district health officials consider to be resilience interventions. In this study, resilience was defined as the capacity of people and systems to mitigate, adapt to, recover, and learn from shocks (Disease epidemics) in a manner that reduces vulnerability and increases well-being (17–19).

The Albertine region suffers recurrent floods and landslides due to heavy rains. These have far reaching impacts to the affected areas escalating recurrent epidemics of cholera. Before cholera vaccination, reports from the ministry of health epidemiology and surveillance department indicated that since the beginning of 2013 the cumulative number of cases reported from the cholera affected Hoima, Nebbi, and Buliisa districts had reached 216 cases and 7 deaths and overall case fatality rate Nationally from these districts stood at 3.2% (20). The region was also at risk for Dysentery and other epidemic prone diseases (20). In 2018, Uganda adopted the oral cholera vaccine (OCV) as part of its integrated strategy to prevent and control cholera, particularly in high-risk areas such as fishing communities, urban informal settlements, and refugee-hosting districts (21). Mass vaccination campaigns, supported by the Ministry of Health, WHO, and partners such as Gavi and UNICEF, at the beginning of the program were rolled out in hotspots including Hoima, Kasese, Kampala, and refugee settlements in West Nile (22). The use of OCV was introduced to complement other interventions such as improved access to safe water, sanitation, and hygiene (WASH).

According to Department for International Development (DFID) practice paper 2011, resilience measurement can be understood from four key elements. These include the context which focuses on ‘resilience of what (systems and processes); disturbance e.g. natural hazard which focuses on resilience to what (shocks and stresses); Capacity to deal with disturbance (Adaptive capacity) and Reaction to disturbance in form of Survival, coping, recovery, learning and transformation (23). Resilience measurement considers context and allows a rational answer to the question of resilience of what’ (24). Once the system or process of interest is determined, the next stage in resilience measurement is to understand the disturbances faced, addressing the question ‘resilience to what?’. In the context of this study, we explored resilience to the cholera outbreak. These disturbances usually take two forms either as shocks (sudden events that impact the vulnerability of the system and process) or stresses (long-term trends that undermine the potential of a system or process and increase the vulnerability of actors within it. According to (23) “the ability of the system or process to deal with the shock or stress is based on the levels of exposure, the levels of sensitivity and adaptive capacities”(23). Adaptive capacities within the context of resilience analysis allow actors to anticipate, plan, react to, and learn from shocks or stresses. (23) The reaction to a shock or stress might be a ‘bounce back better’ for the system where capacities are enhanced or sensitivities and exposures are reduced, leaving a system more able to deal with future shocks and stresses (24). On the other hand, an alternative reaction might be a ‘bounce back’ to a normal, pre-existing condition, or to ‘recover but worse than before’ resulting in reduced capacities (24). The worst-case scenario is where a system might not bounce back at all, but ‘collapse’, leading to a catastrophic reduction in capacity to cope in the future (23)

The study assessed the resilience of the health system to the effects of cholera outbreak with a goal of informing the design interventions that strengthen the resilience of the health system, The findings from the study will inform the systems thinking workshop that will convene a team of stakeholders involved in health care delivery system and facilitate them through a systems thinking process. The workshop will focus on the relations between the components of the resilient system, events, interactions and feedback between these components. Understanding the interrelationships between the different components of a resilient health system will be a first step in designing the conceptual framework for strengthening the health system in the event of disasters. This paper focused on the identified resilience themes as a basis for better understanding of health system resilience, especially in handling emergencies like cholera. In the follow-on paper, the systems interventions generated through the systems thinking workshop will be published. This study therefore explored resilience at the local government, community levels understanding the adaptive mechanisms implemented to prevent cholera and strengthen the health system.

## Methods

### Study design and study area

A qualitative study was conducted in the Albertine districts Hoima, Kasese and Kikuube in Western Uganda. The Albertine region was selected because it suffers recurrent floods and landslides due to heavy rains with far-reaching impacts on the affected areas, escalating recurrent epidemics of cholera, malaria, and viral hemorrhagic fevers. In Kasese district, Mpondwe – Lhubiriha Town Council and Nyakiyumbu sub county were selected. In Hoima district, Kigorobya and Buresuka sub counties were selected while in Kikuube district, Kabyooya and Kyangwali sub counties were selected. Specifically, Focus Group Discussions (FGDs), Key Informant Interviews (KIIs), and In-Depth Interviews (IDIs) were used in this study as these methods are ideal capturing perspectives from multiple levels of the system. The FGDs explored community experiences, behaviors, and perceptions of the health system’s response, revealing common coping strategies. The KIIs provided expert insights from district level health officials, and administrators, highlighting systemic strengths, gaps, and preparedness measures. While IDIs allowed detailed exploration of personal experiences from patients that suffered cholera as well as the caregivers uncovering operational realities and sensitive issues that may not emerge in group settings. The use of these qualitative methods provided a holistic, context-specific understanding of health system resilience, allowing for triangulation of data and in-depth analysis of the multifaceted factors affecting cholera response in the study districts.

### Study Participants

Key informant interviews with District Health Officers (DHOs) or Assistant DHOs in charge of Environmental Health, Health Inspectors or Water Officers Health Facility In-Charges, representatives from NGOs or CBOs working on water, sanitation, or hygiene and Surveillance Focal Person were conducted. Key informants were selected purposively based on their knowledge, position, and experience in cholera response including the understating of the historical perspective. To be included in the study, the key informant must have been either the current or previously worked within the district of the study in the period between 2010 to the time of the study (2025). Participants must have participated in at least one cholera response and must have consented to be interviewed. Focus Group Discussions (FGDs) participants were selected based on whether the participants were at-risk or affected by cholera, caregivers of patients who previously suffered cholera, a representative of health workers serving at the community level, a village health team member, community leaders and influencers as well as vulnerable groups like refugees where applicable. On the other hand, in-depth interviews were structured conversations with individuals who themselves survived cholera (survivors) or the caregivers of patients who suffered cholera.

### Data collection procedures

Prior to data collection, trained and experienced researchers with a minimum of a bachelor’s degree and previous experience in qualitative research were selected. The research Assistants were selected from the research assistants pool of the Makerere University School of Public Health (MakSPH). The MakSPH over years has established research assistants pool built through different research projects with language profiles. In the initial process, 15 research assistants for greater Hoima and ten (10) for Kasese were selected from the pool and invited for interviews. Six (6) for the greater Hoima and four (4) for Kasese were selected and later trained by author1 for 4 days of which one (1) day was for the pr-test of the study tools in Makerere 3 parish-a place that has over years been hit by cholera outbreak. Following the training, they were dispatched for data collection. In each of the study districts, team of the research assistants headed by author1 begun with an introductory meeting with the District Administrator (the Chief Administrative Officer), District Health Officer, and the Resident District Commissioner. In the introductory meetings, author 1 introduced the study goal, the study sites and the categories of the respondents. The meeting with the District Health Officers also supported in the identification of the Key informants and the Health inspectors at the sub county level. The District Health officers also provided contacts of the key informants and in some cases, made interview appointments. Using the contacts of the health inspectors provided by the District Health Officers, the study team contacted them to schedule the identify the Focus group discussion participants as per the categories described, identify the venues for conducting the FGDs as well as contacting and scheduling interviews with in-depth interview participants. At each sub county, schedules for FGDs and IDIs were shared with the study team who then implemented as communicated by the health inspectors. In each sub county, 2-3 FGDs were conducted. The table 1 below shows the distribution of the FGDs, KIIs and IDIs is included below

**Table 1:** Number of FGDs, IDIs and Key Informant Interviews by district.

| District | Study areas (sub counties) | FGDs | KIIs | IDIs |
| --- | --- | --- | --- | --- |
| Hoima | Buseruka & Kigorobyia | 2 | 4 | 3 |
| Kikuube | Kyangwali & Kabwoya | 3 | 4 | 3 |
| Kasese | Mpondwe Lhubiriha Town Council & Nyakiyumbu | 5 | 5 | 3 |

All IDIs were conducted at the household level whereby the data collection teams (An interviewer and a notetaker) visited the household to conduct each interview. For the Key Informant Interviews, they conducted in district offices and Health facilities for the District officials and Health Facility in-charges respectively. For the FGDs, they were either conducted at the Health facility or within the community as guided by the Health inspectors. All FGD participants were provided with transport refund worth Uganda Shillings 20,000 (approximately $ 5) as approved by the Ethical review Board. All interviews were held during the day between 8am and 7pm and each interview (FGD, KII & IDI) were conducted by two research assistants (interviewer/facilitator and note taker). Verbal consent was sought from all the participants after explaining to them the study objectives, risks and benefits. Using translated interview guides, they moderated the discussions while the note-takers captured detailed field notes in their notebooks. Audio recorders were also used to supplement the field notes captured by the note-takers. The languages used for the translated guides were in the local dialects of *Runyoro/or Rukiga (for Banyoro or Bakiga), Kiswahili (for Lugbhara community)* in Hoima and Kikuube (Greater Hoima) and *Lhukonzo* for Kasese. On average, each FGD took 90 minutes while IDIs and KIIs took 60 minutes.

### Data collection tools and variables

The interview guides were used for both (IDI, FGDs and KIIs. The guides captured generatic resilience variables including 1) Health management capacities (Health system preparedness, alert systems, interdependence, coping mechanism, coordination, financing, reporting and legitimacy 2) Resilience (Absorptive capacity, Adaptive capacity, Transformative capacity and adaptive mechanisms. Five themes were explored as detailed in table 2 below

**Tabe 2:**
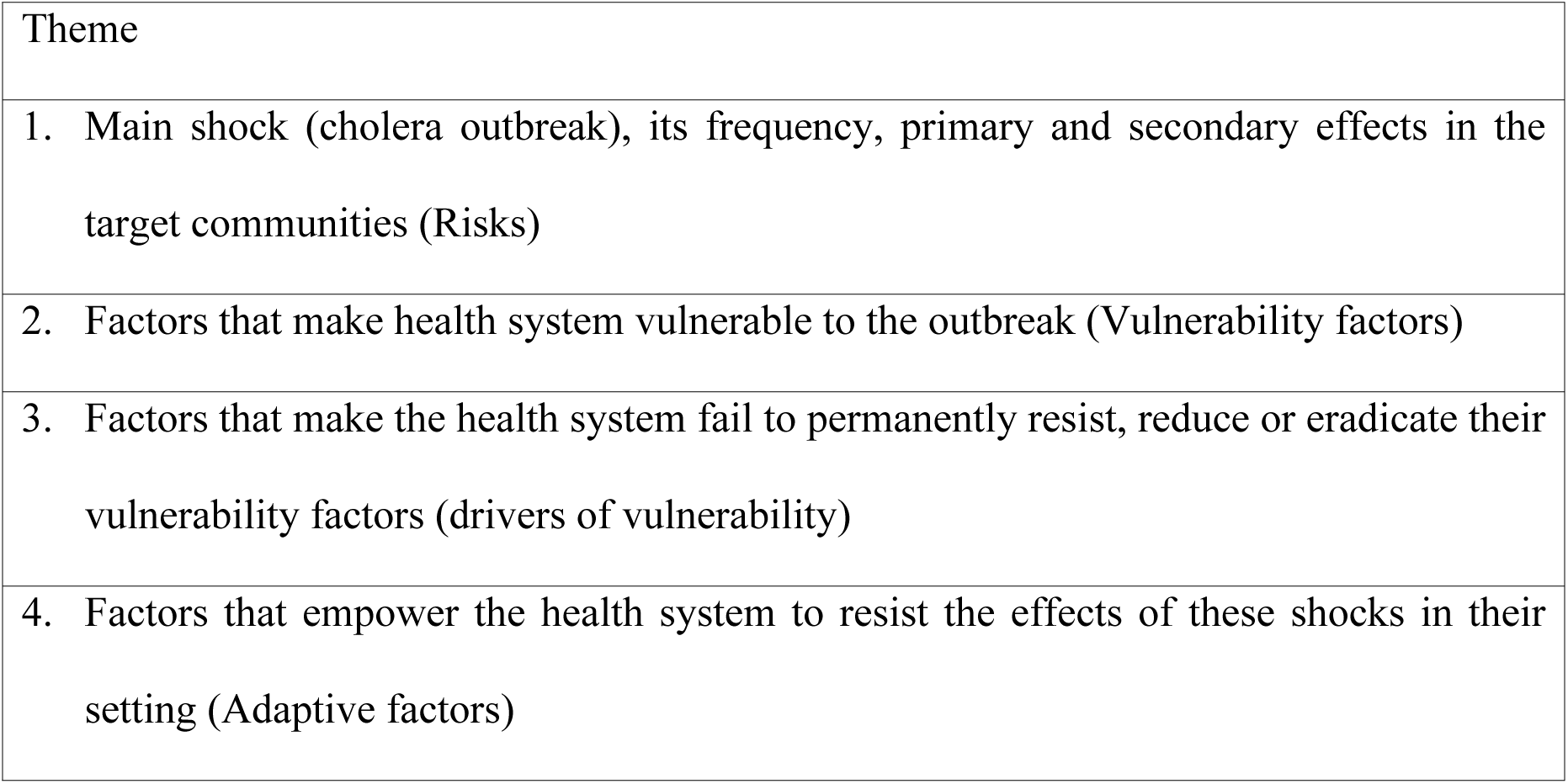
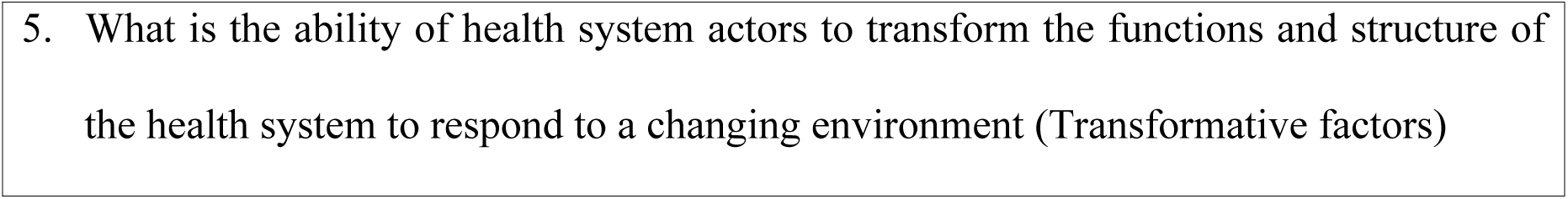
General themes that were captured in the interview guides during data collection.

In this study, subjectivity was addressed through triangulation of data collection methods (FGDs, KIIs, and IDIs), reflexivity of the research team to explore beyond the interview guides by probing. To maintain rigor and achieve phenomenological saturation, the data collection team encouraged active participation especially during the FGDs as well as asking through rapport-building with study participants, asking open-ended questions during the interactions, and the use of probes to elicit deeper insights and explore emerging issues beyond the initial interview guides. Evey evening, author 1 who participated in the data collection in all study districts would hold debrief meetings with the research assistants and listed to selected recordings as part of maintaining an audit trail and ensuring that the data provided rich and contextualized descriptions, accurate reflection of participants’ experiences and the complexity of the health system’s response to cholera.

### Data analysis

The audio tape recordings were transcribed verbatim by selected research assistants fluent in the local languages and with prior experience in transcription. The authors (NT, RWM & AML) developed a coding scheme following multiple readings of the transcripts and field notes. Out of the 10 research assistants that collected the data, three were with prior experience in data analysis were selected to support data analysis. Using latent content analysis, the identified codes were arranged into categories and emerging themes using excel. Analysis was done in two stages, first, the manifest content analysis followed by latent content analysis. Transcripts generated from the transcripts were read and re-read by three 3 selected research assistants, who then assigned codes (Open coding). Data meaning units were then aligned under their respective codes. This was followed by axial and selective coding to develop higher codes and sub-themes. Sub-themes were reviewed further to develop overarching themes (typologies). Typologies were the basis for deriving constructs in the resilience assessment for the health system. The overarching themes were verified by NT and RWM. The key adaptive mechanisms employed by the health system within the study districts (resilience factors) were captured accordingly as themes. Some transcripts were analyzed with Artificial Intelligence using Chatgpt4.5. In the Chatgpt chatbot, an excel with transcript statements was uploaded and commanded as follows “Use the attached excel and analyze to fill columns on the condensed meaning of each statement, generate a Interpretation of the underlying meaning from the same statement, a sub dimension and Dimension. Fill all rows in the Excel file with condensed meaning, interpretations, sub-dimensions, and dimensions”. The generated information was then re-read by NT and the research assistants ensuring alignment.

## Ethical considerations

Ethical approval was obtained from the Makerere University School of Public Health Research and Ethics Committee (SPH-2024-675). Prior to any of the interviews, including group discussions, a consent form was read out to the participants who were given a chance to ask questions. Approval was also sought from district leaders who were informed about the study before the data collection. Issues of anonymity and confidentiality were emphasised and FGD participants were assigned numbers to conceal participants’ names and identifications

## Results

A total of eighty-seven (87) people participated in the study 46 men and 41 women, respectively, with an average of 9 people per FGD. A total of nine (9) In-depth, ten (10) FGDs and thirteen Key informant interviews were conducted. The majority of participants were married and had completed primary education. Seven key themes were identified as drivers of resilience to cholera outbreak: Epidemic Preparedness and Response Planning; Health Education and Behaviour Change Communication; Water and sanitation; rapid response, treatment and isolation; Community Mobilization and VHT Networks; community and cross-border surveillance. These emerging themes are further elucidated in this section.

## Oral vaccination of cholera

Participants said that cholera vaccination campaigns, particularly effective in 2018 following an emergency OVC campaign, contributed significantly to outbreak reduction in the subsequent years. Respondents from Key Informant interviews, Focus group discussions, and in-depth interviews emphasized the overall positive impact of the vaccination initiative, noting a marked decline in cholera cases post-vaccination. In the Focus Group Discussions (FGDs), community members acknowledged that cholera vaccination had been introduced and that many had received at least one dose. They perceived that since vaccination, cholera cases had reduced, although some remained uncertain whether the decline was solely due to vaccination or other interventions. However, in the In-Depth Interviews with cholera survivors in Kasese and Kikuube, the respondents stressed the importance of preventive health education and improved sanitation but confirmed that vaccination, along with hygiene measures and provision of safe water, was essential to reducing community vulnerability. This survivors’ account underscored how vaccination works best when complemented with community sensitization and household-level hygiene practices. Key Informant Interviews (KII) with district health officials also confirmed that, vaccination was highlighted as a government-led intervention, with the official noting that while acceptance was generally high, a few individuals resisted vaccination on cultural or religious grounds, such as Jehovah’s Witnesses. The official also emphasized the need for consistent supply and distribution, since vaccines are only delivered periodically and may not always reach lower-level facilities.

> *“The vaccination, yes. We did the cholera vaccination… one of the contributing factors why we no longer have cholera.” KII Kasese*
>
> *“Since that time of vaccination, cholera has stopped and we have not had another outbreak in Kyangwali sub-county”IDI, cholera survivor Kikuube*

Key informants in Hoima stressed that persistent education and local leadership intervention significantly improved vaccine acceptance rates. They reported that vaccine coverage was targeted strategically in outbreak-prone areas (sub counties) especially those along Lake shores and in the refugee settlements and host communities, achieving substantial community immunity which helped prevent subsequent outbreaks with coverage reported to be above 80% as some community members refused to be vaccinated while others missed out due to the mobile nature of the activities.

> *“People don’t refuse vaccination. It is few people who refuse… Others have their own culture. Others, religion. Like Jehovah’s Witness, they can’t vaccinate their children.” KII Frontline worker Kasese district*

### Epidemic Preparedness and Response Planning

In Hoima, Kasese and Kikuube, there were concurrencies in the FGDs on how participants described preparedness which was mainly through community-level engagement, noting the mobilization of Village Health Teams (VHTs) who were deployed to sensitize households on hygiene and safe water practices. The participants emphasized that such grassroots preparedness was critical in reducing cases especially during active outbreaks. Meanwhile, the In-Depth Interview (IDI) participants reflected an experiential perspective, highlighting how timely intervention related to treatment by government and community sensitization prevented further deaths once treatment teams and preventive messages were deployed. The survivors stressed that preparedness should not only mean medical response but also proactive education and community empowerment before outbreaks occur. From the KIIs, epidemic preparedness was viewed from a systems perspective, focusing on the district’s ability to mobilize resources (personnel, essential medicines), rapid establishment of treatment centers, and coordinate with stakeholders, including Ministry of Health, VHTs, Non-Governmental Organizations (NGOs) and local leadership.

Interviews with the District Health officers from the three districts confirmed the availability of trained Teams and Structures and the districts have structured Integrated Disease Surveillance and Response (IDSR) systems mainly at some HCIII and above, ensuring preparedness at multiple administrative levels (Village-parish-subcounty and district) coupled with a pool of specialized trained personnel at Clinical Officer level capable of rapid deployment. In Kasese district, it was reported by District Health Office that majority of these specialized trainees are based at Bwera Hospital while in the greater Hoima, they were distributed across different facilities with Kyangwali HCIV hosting the majority due to being a refugee host community. The capacity of the districts was reported to be strengthened through the establishment of temporary isolation facilities established quickly during outbreaks with some being upgraded to support future outbreaks. In Kasese, an isolation facility which was established at Bwera Hospital has been maintained while in the greater Hoima, it was reported that an isolation and treatment center has been established at the Hoima Regional Referral Hospital, at Kikuube Health Center IV and Kyangwali Health center IV in Kikuube district.

The district health officials and representatives of NGOs interviewed acknowledged gaps, particularly the lack of regular training for health workers at lower levels especially Health center IIs & IIIs, insufficient medical supplies and emergency kits and limited infrastructure, which constrained effective preparedness and timely response. Collectively, these perspectives revealed epidemic preparedness and response as a combination of community mobilization, health system capacity, and individual experiences of timely intervention in seeking care and implementing household level remedies.

Relatedly, officials in the three districts reported the availability of epidemic preparedness and response plans specific to cholera which were developed and promptly activated during outbreak alerts. They reported that the response plans contain key elements like the establishment of the district cholera task force expected to have regular meetings, training of the rapid response team, skilled health workers deployment plans, water and sanitation facilities requirements, behaviour change communication mechanisms, health Education and support supervision mechanisms.

> *“The Ministry…we have them submitted, but you find other things happening when the outbreak has ended. For example, they are now beginning to bring the IC materials to teach people. When the outbreak has ended, they will tell you no funding.” KII Kikuube*
>
> *“We normally develop epidemic preparedness and response plans, depending on the outbreak that comes. When an outbreak comes, we have a response plan in place and we swing into ground.” KII Kasese*

### Health Education and Behaviour Change Communication

During interviews with key informants and as confirmed by the FGD participants, all participating districts conducted education campaigns on cholera transmission modes and prevention methods both during active outbreaks and in routine health education when there are no active outbreaks. In the Focus Group Discussion (FGD) at Katholhu Village Kasese Disttrict, participants emphasized that health education was a key preventive measure, citing how VHTs and health workers regularly visited communities to teach about food hygiene, boiling water, and latrine use. The FGD participants at Kibiro village in Hoima district noted that such consistent messaging gradually influenced behaviour change, with fewer people engaging in risky practices. From the Key Informant Interview (KII) with the a representative of an NGOs in Kikuube district, health education and behaviour change communication were framed as structured interventions led by the district, often in partnership with NGOs and local leaders with VHTs at the centre of implementation. The district official in Hoima highlighted the use of community dialogues, schools, churches, and local council platforms as avenues for communication of preventive and treatment related messages. Relatedly, the In-Depth Interview (IDI) with a cholera survivor in Kasese reflected the personal impact of health education, with the respondent recounting how messages about safe water and food hygiene helped families adopt safer practices after experiencing the outbreak. This perspective from the survivor underscored the effectiveness of behaviour change communication when it is practical, relatable, and directly linked to lived experiences.

> *“We trained about 5 VHTs per village. After training, they started home-to-home visiting. The number of patients decreased significantly.” KII Kasese*
>
> *“Due to intensive health education… they came to understand the cause of the disease and why it was spreading” KII Kikuube*
>
> *“From the time the health workers and government sensitized us, since then we have not experienced cholera outbreaks in our community.” – FGD, Hoima*
>
> *“People knowing that cholera is dangerous, people learning the transmission…people were heavily educated in the mode of spread, and now people know that it is associated with the faeces.” KII Kasese*
>
> *“You would find everyone was now boiling water, covering their food, and eating hot food… People change behaviour because they know. So the change in behaviour alone makes the disease disappear.” KII Hoima*

### Water and Sanitation

In the Focus Group Discussion (FGD) at Mpondwe Lhubiriha Town Council Kasese District, participants described how the provision of safe water sources, such as boreholes, alongside health education on hygiene, significantly reduced cholera cases within weeks of the previous outbreaks. The Kyangwali FGD participants acknowledged community-driven measures like banning roadside food vending, and sale of cooked fish at the landing sites which complemented sanitation efforts to reduce on cholera outbreaks. In the In-Depth Interview (IDI) with a caretaker cholera survivor in Kikuube, a responded highlighted how lack of access to safe water and sanitation facilities especially I villages near the lake initially left households vulnerable, but the installation of boreholes and community sensitization brought tangible improvements. Another IDI respondent in Kyangwali Kikuube district noted a direct linkage between receiving safe water interventions in their community and a noticeable decline in cholera outbreaks overtime. From the Key Informant Interview (KII) with the Assistant District Health Officer in Kasese, water and sanitation challenges were framed in terms of persistent open defecation, seasonal settlements in farms lacking latrines, and unsafe water sources that sustained cholera transmission. While in Hoima, the seasonal economic activities were mainly around fishing with mobile fish mongers lacking latrines at their lake shore established temporally settlements. The officials in both Hoima and Kasese emphasized the need for infrastructural investment and stronger enforcement of sanitation bylaws.

> *“The VHTs always passed around the area… trained on food and food hygiene… and now that act is decreasing.” FGD Kasese*
>
> *“Most people didn’t have toilets. They used to drink unboiled water and they used to take roadside foods… during seasons, many people come from other areas… and they*
>
> *don’t have latrines. This practice has changed over time as all households have been compelled to have pit latrines and roadside vending of food was banned.” KII Kikuube*
>
> *“Now there was an increase in clean water supply…they started protecting water sources and people were learning to use safe water, it could be another reason why we have no virus.” IDI caretaker of cholera survivor Kikuube*

### Rapid response, treatment and isolation

In the Focus Group Discussion (FGD) at Nyakiyumbu Village and Mpondwe Lhubiriha Town Council Kasese district, community members highlighted the establishment of treatment centers at Bwera Hospital and later at Katholhu Health Center II as pivotal in reducing cholera deaths. They described how once treatment teams arrived and facilities were organized, mortality dropped drastically compared to the initial stages of the outbreak before the isolation and treatment centres were established. The Key Informant Interviews with district officials in Kasese and Kikuube emphasized the districts’ role in coordinating partners like, mobilizing health workers, and ensuring supplies of antibiotics and rehydration solutions. It was reported that during the active outbreaks, Health centers received support in form of supplies such as IV fluids and ORS. For example, it was reported that since 2017 in Hoima district, health team established an isolation center big enough to segregate patients by gender. The officials confirmed that during active outbreaks, support was mainly provided by the district, Ministry of Health, and United Nations International Children’s Emergency Fund (UNICEF). The officials also acknowledged logistical constraints, particularly limited staff and delayed establishment of treatment sites, which sometimes allowed outbreaks to spread further before control was achieved. The interviews with two cholera survivors in Hoima and Kasese recounted how before health workers intervened, several deaths occurred, but once treatment and isolation were provided, survival improved dramatically. These insights from the survivors demonstrated the role of survivors’ experiences in shifting community perceptions moving from attributing illness to witchcraft to recognizing it as a treatable disease, thus increasing acceptance of medical interventions.

> *“In Kasese, we have at least six responders. In case an outbreak happens, the Kasese team is able to move because the team is built with capacity to handle any outbreaks.”*
>
> *KII Kasese*
>
> *“Though we don’t have a permanent isolation units, we have temporary units in all facilities designated to manage outbreaks of such nature.”KII Hoima*
>
> *“We opened up a treatment center at Bwera Hospital… we managed 291 patients but got only one death.” — KII Frontline Health Worker, Kasese*

### Community Mobilization and VHT Networks

Community Mobilization emerged as a central pillar in cholera control across Kasese, Hoima and Kikube. In the Focus Group Discussions (FGDs), community members repeatedly emphasized the visibility of Village Health Teams (VHTs) during outbreaks. They described how VHTs and local leaders convened meetings, conducted household visits, and delivered health education on safe water, food hygiene, and sanitation practices. Participants emphasized that the VHTs’ efforts were often credited with shifting behaviors, such as discouraging roadside food vending and promoting latrine use, which directly reduced cholera cases and outbreaks. Similarly, Key Informant Interviews (KIIs) with district officials underscored VHTs as the frontline link between the health system and the community. Officials highlighted VHTs mobilization for oral cholera vaccination (OCV) campaigns, their role in case detection and referrals, and their ability to rally local structures, including churches and local councils, for coordinated prevention. However, they also noted challenges such as limited training, supervision, and facilitation, which constrained the full potential of VHTs in sustaining community mobilization. From the In-Depth Interviews (IDIs) with survivors, VHTs were reported to be often the first to arrive in homes with preventive messages and were trusted sources of guidance. Survivors described how VHTs reassured families, corrected misconceptions about cholera being caused by witchcraft, and encouraged early care-seeking at treatment centers.

> *“We have trained structures… beginning with the VHT structures regularly trained in the integrated disease surveillance and response.”KII Hoima*
>
> *“When we get a challenge in every village, and this information reaches the facility through the network of VHTs, the facility will communicate a response through the same network.”KII Frontline Health worker, Kikuube*
>
> *“The households through the support of community health workers… have a very big role in mobilising themselves against such pandemics.”KII Kasese*

### Community and cross-border surveillance

Community-based surveillance played a crucial role in detecting and responding to cholera outbreaks at the grassroots level. From the Focus Group Discussions (FGDs), participants explained how Village Health Teams (VHTs) and local leaders acted as the “eyes and ears” of the health system, quickly identifying suspected cases and reporting them to health facilities. Participants reported that they valued this system because it enabled faster response and reduced delays that often cost lives. They reported that Health talks by both VHTs and Health workers, home visits, and reporting by VHTs created a continuous flow of information between households and the formal health system. From the Key Informant Interviews (KIIs), district officials highlighted the importance of structured surveillance networks, and confirmed that VHTs submitted weekly reports during active outbreaks, district epidemic preparedness committees in both districts were reported to be active, and coordination with partners through quarterly meetings were reported to have been held in all the three districts. However, district officials acknowledged weaknesses, such as under-reporting, lack of training for some VHTs, and limited resources for follow-up, which sometimes undermined the efficiency of community surveillance.

On the other hand, cross-border surveillance introduced unique complexities. In the FGDs, community members observed that population movement between Uganda and the Democratic Republic of Congo (DRC) was common, especially through informal crossings, increasing cholera risk. They noted that surveillance efforts were sometimes disrupted by the porous nature of the border. The KIIs described efforts by district leaders to hold joint meetings with Congolese counterparts during outbreaks of cholera, Ebola, and even COVID-19, where prevention strategies and community sensitization were coordinated. They reported that these meetings strengthened cross-border communication but were irregular and heavily dependent on available funding. From the In-Depth Interviews (IDIs), they perceived that outbreaks often began with cases linked to cross-border traders or travelers, reinforcing the community belief that movement across the border was a key driver of infection.

> *“Our surveillance teams here should freely be allowed to interact with the surveillance team across and they should be able to share cases and the situation freely.”KII Kasese*
>
> *“The coordinated response is there by communication. But in responding quickly, sometimes the constraint is delayed by resources.”KII Frontline Healthworker, Hoima*
>
> *“Much of the origin of the cases we get… the primary case, call it the index cases, are always from Congo, so our surveillance must take into consideration issues across the border.” KII Hoima*

## Discussion

In this study, seven (7) major resilience factors were identified, including Oral vaccination of cholera; Epidemic Preparedness and Response Planning; Health Education and Behaviour Change Communication; Water and Sanitation; Rapid response treatment and isolation; Community Mobilization and VHT Networks; and Community and cross-border surveillance.

Historically, cholera, a disease associated with poverty (25) has posed a significant health challenge to the Albertine region since 1970s. The disease has caused both psychological, social and economic impacts on the households, communities and the health system. As such, the affected systems have overtime instituted different interventions that have strengthened resilience at various levels. The interventions implemented at the household, community as well as health system have contributed to the declining trend of cholera cases with no outbreaks reported since 2019 an indication that Uganda might be moving towards heightened resilience towards water and sanitation diseases. It is also important to underscore the role of these interventions in broader control and prevention of disease outbreaks and health system strengthening.

As part of the health system strengthening towards cholera prevention and control, Uganda developed Operational Guidelines for the National and District Health Workers & Planners that emphasizes critical elements in cholera control are Prevention, Preparedness, Response and an efficient surveillance system (26). These elements are implemented in an integrated manner. These elements speak to resilience where health system capacities are enhanced, exposures are reduced, leaving a system more able to deal with future shocks and stresses (24). This framework is followed by a robust package of interventions ranging from cross-border surveillance, preparation of district and national plans of action during and after the outbreaks, mass vaccinations, to WASH interventions implemented at the household and community level

The findings indicate that the introduction of oral cholera vaccines (OCV) is a critical intervention that has significantly reduced cholera incidence in the study districts. Uganda introduced the cholera vaccine in 2018 with over 3M doses administered in 13 hotspot districts during 2018–2020 for both preventive and reactive campaigns (27). The reported vaccination coverage was between 78% and 90% (28). The endemic districts were targeted for vaccination including the greater Hoima (Hoima City, Hoima district and Kikuube district) and Kasese. It is important to note that WHO recommended oral cholera vaccines, combined with sustained sanitation improvements and hygiene practices, as significant measures to reduce cholera incidence (19). Relatedly, studies conducted in Haiti, Bangladesh, and other cholera-endemic regions corroborate these findings, suggesting high vaccine efficacy in outbreak prevention and control, especially when accompanied by WASH interventions(29, 30). The findings imply that integrating Oral Cholera Vaccination (OCV) into Uganda’s cholera control strategy should be institutionalized as a routine and sustained intervention in hotspot districts, rather than as a one-off campaign. While OCV has demonstrated significant impact in reducing incidence, its long-term effectiveness depends on systematic booster campaigns, expanded geographic coverage, and integration with WASH programs. Therefore, programmatically, the Ministry of Health and partners should institutionalize OCV in national immunization schedules for high-risk populations in hotspot districts and plan periodic booster doses (every 3–5 years) to sustain immunity and prevent resurgence. Similarly, there is need to strengthen integration of OCV with WASH interventions, ensuring safe water supply at community level, sanitation infrastructure, and hygiene promotion accompany vaccination.

Cholera Vaccination campaigns were accompanied by other interventions such as improved water, sanitation, and hygiene (WASH) practices, health education, and behavior change initiatives. For instance, Health education, provision and use of protected water sources significantly supported the sustainability of cholera reduction post-vaccination. These interventions are well documented as critical to the elimination of cholera disease. What emerges from these findings is not only that WASH, health education, and vaccination were implemented, but that these interventions appear to be shifting cultural practices and community norms. For example reported practices where communities began to routinely boil water, use latrines, and avoid unsafe food vending, practices that previously were somehow resisted suggesting a cultural shift where cholera-preventive behaviors are moving from being emergency responses to being part of everyday social norms. Similarly, there was a strong involvement of VHTs, local councils, and religious leaders in the cholera prevention and treatment intervention which reflects a deeper social embedding of cholera prevention into existing community structures. Such ownership makes interventions more resilient than externally driven campaigns. In a study by Gabutti (25), it was clearly documented that in developing countries, cholera transmission can be effectively prevented by enhancing access to clean water, proper sanitation, and improved hygiene practices, alongside administering oral cholera vaccines (OCVs). In addition, the study recommended that travellers especially those travelling to endemic countries can reduce their risk by consistently practicing hand hygiene and ensuring the safety of the food and water they consume (25).

While Uganda’s health system resilience to the cholera outbreak has been strengthened, there are a number of critical issues that need to be addressed if this positive trend is to be sustained. Issues around sanitation infrastructure, particularly in transient farming and fishing communities, exacerbates open defecation and hygiene problems. Access to safe drinking water remains insufficient, prompting reliance on unsafe sources like lakes and rivers. Behavioural challenges persist; despite temporary improvements during COVID-19, practices like handwashing have declined post-pandemic. Since the development of sanitation infrastructure falls outside the purview of health issues, we recommend that institutions responsible for urban planning, sanitation in local government, and the Ministry of Water and Environment should working in consultation with ministry of Health address these issues given the requisite expertise and jurisdiction. Health facilities often have limited resources, trained personnel, and effective infection prevention measures (31, 32). High population mobility, especially cross-border movements with the Democratic Republic of Congo, complicates surveillance and control efforts. Furthermore, weak inter-sectoral coordination that does not involve the cross-border districts in DRC undermines integrated sanitation and hygiene interventions (33, 34).

## Conclusion

To sustain the resilience of the system to a cholera outbreak, Periodic Vaccination Campaigns especially targeting transient and high-risk groups should be implemented. Our study gives evidence that since the vaccination in 2018-2020 that did not cover 100%, there has not been any other vaccination targeting those who missed out during the initial exercise. It would be critical for the country to consider booster doses are advised in every 3 to 5 years, especially in the districts where OVC was conducted. Additionally, continuous adherence to Hygiene Practices such as maintaining behaviors related to boiling water, handwashing, and using latrines to complement vaccine-induced immunity is critical. Using community health structures like VHTs, regular campaigns using local languages and community influencers should target messages on WASH as well as enforcing sanitation regulations.

## Author Contributions

Conceived and designed the study: NT&RWM. Analyzed the data: NT. Contributed analysis tools: NT,AML, RWM, SK, FS, GB. Wrote the first draft paper: NT. All authors reviewed the paper and approved the submission

## Data Availability

I can avail the excel sheets that i used for anylysis and conding if required. I can also avail the transcripts from the interviews conducted if required

## Notes

### Competing Interest Statement

The authors have declared no competing interest.

### Funding Statement

This study is fully sponsored by myself as part of my PhD studies and there is no individual or institution that contributed funds towards this research

### Author Declarations

Makerere University School of Public Health Research and Ethics Committee (SPHREC)

